# The difference method approach for sampling order constrained parameters: an improved implementation and important limitations

**DOI:** 10.1101/2023.06.12.23291285

**Authors:** Daniel Hill-McManus

## Abstract

A health economic model may include a set of related inputs whose true values are uncertain, but that can be assumed to follow a logical order. Various approaches are available for performing probabilistic sensitivity analysis while preserving the order constraint, one such approach is known as the difference method. The difference method approach appears to have many of the required properties, has been endorsed by good practice guidelines, and is likely to prove a popular approach. However, the proposed implementation of the difference method approach is cumbersome, requiring numerical estimation, which might present a barrier to its adoption. Furthermore, it is unclear whether the method can always be applied to 3 or more model inputs and whether it is unbiased across all possible input values. This study has investigated these three issues for ordered inputs bounded between 0 and 1. An analytic solution is given that allows for more straightforward and compact implementation. The difference method approach cannot always be applied to a set of 3 or more model inputs, and this depends on the relative size of the variances of the logit transformed Beta distributions fitted to each variable. The approach can also produce samples with biased means and variances under certain combinations of input means and variances. It is recommended that the difference method approach be considered, however, an understanding of its limitations is necessary in order to identify such cases.

## 1. Introduction

Health economic decision models routinely use sensitivity analysis to assess the extent of uncertainty in model outcomes arising due to uncertainty in model inputs. Probabilistic sensitivity analysis (PSA) is used to estimate the uncertainty in the outcomes of a decision model due to the uncertainty across the joint distribution of all inputs [1]. This provides a sample of model outcomes via Monte Carlo simulation from which desired summaries can be obtained, such as the mean health effects or mean costs. Summaries obtained from such samples should typically form the basis for decision making where decision models are non-linear [2].

It is not uncommon for there to be related subsets of the model inputs whose true values are uncertain, but whose ordering can be assumed known. The utility values describing the quality of life for a set of health states in a cohort model may be a common example. Where the health states are defined according to the severity of a condition which impacts quality of life, it may be reasonable to assume that it is not possible for a more severe state to be associated with a higher utility than a less severe state. Ignoring this order constraint, and sampling independently during PSA, may then result in realisations of the model inputs in which the logical ordering is reversed. These samples could, therefore, imply a treatment that reduces disease severity leads to lower quality of life.

Ren et al. have proposed a potential solution to this problem, known as the difference method (DM) approach [3]. Their study described three criteria that any method for ordered sampling should satisfy: 1) it produces samples of the inputs within the appropriate bounds, 2) it produces samples of the inputs with the desired summary statistics, and 3) it produces samples of the inputs with a plausible emergent correlation structure. It is then demonstrated via a simulation exercise that the DM approach meets these criteria, whereas the main alternatives; independent sampling, modified independent sampling and the common random number generator approaches, do not.

An ISPOR Good Practices for Outcomes Research Task Force Report [4] recommends use of the DM approach to avoid sampling inconsistent health state utility values. The recent NICE health technology evaluations methods guide [2] also refers to sampling of ordered inputs, and suggests considering approaches that “neither artificially restrict distributions nor impose an unsupported assumption of perfect correlation”. Therefore, there is likely to be an increasing interest and application of the DM approach within health economic models.

To support wider adoption and appropriate application of the DM approach, there are three issues that would benefit from further study. The first is whether the current approach, which requires numerical approximations prior to generating samples, can be improved to obtain a simpler more compact implementation. The second is how, and whether it is always possible, to extend the DM approach to more than two input variables. The third issue is whether the DM approach can be expected to provide unbiased samples (objective 2 above) for all possible input variable means and variances.

The aim of this study was to further develop the DM approach and to provide insights into the issues outline above. An analytical solution to the previous simulation approach is provided to simplify the implementation of the DM approach, particularly if using a spreadsheet type software for decision modelling (an Excel template is provided). It is shown how the DM approach can be extended to more than two variables, along with when this will prove possible. Through a simulation exercise, it is found that the DM approach can produce biased sample means and variances. Health economic modellers should be aware of the potential limitations of the DM approach when considering how to handle the issue of ordered sampling.

## 2. Difference method approach summary

Before proceeding with an investigation of potential issues relating to the DM approach, a summary of the approach proposed by Ren et al. for inputs on the 0,1]interval is given below [3]. While Ren et al. described the DM approach for inputs on the interval 0,1]and [0,∞), this study has only focussed on the interval 0,1]. This assumes two input variables *X*_1_ and *X*_*2*_ that are bounded between 0 and 1 for which the means (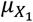and 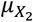) and standard errors (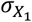 and 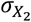) are known. The two inputs are defined such that 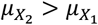. The main steps are as follows:

- In step 1, a Beta distribution is assumed to characterise the uncertainty for each input and the two parameters of each distribution are derived using the means and standard errors. For example, for the first input it can be written 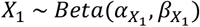.
- In step 2, the inputs are mapped to the real line via a logit transformation such that *X*′_1_ =*logit*(*X*_1_)and *X*′_*2*_=*logit*(*X*_*2*_). The means and variances for *X*′_1_ and *X*′_*2*_ are needed and must be approximated by applying the logit transformation to a large number of samples of *X*_1_ and *X*_*2*_.
- In step 3, a difference distribution is defined to characterise the difference between *X*′_1_ and *X*′_*2*_. A Gamma distribution is proposed, and the shape and rate parameters are calculated to give a distribution with mean and variance equal to the differences between *X*′_*2*_ and 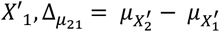 and 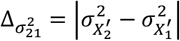. Therefore, it is possible to probabilistically simulate differences for *X*′_*2*_ and *X*′_1_ according to 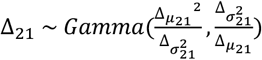.
- Step 4 combines sampling of *X*′_*2*_, *X*′_1_ and *Δ*_*2*1_ with inverse logit transformation to produce the desired samples of *X*_1_ and *X*_*2*_. The algorithm depends on the relative size of the variances of *X*′_1_ and *X*′_*2*_, if 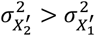then *X*′_1_ samples are used and *X*′_*2*_ obtained as *X*′_1+_*Δ*_*2*1_ However, if 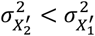then *X*′_*2*_ samples are used and *X*′_1_ obtained as 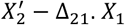 and *X*_*2*_ are finally obtained via inverse logit transformation of 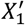and 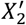.

## 3. Further developments

This study has set out to demonstrate i) a more efficient implementation of the DM approach that does not require numerical approximation of expectations and variances for logit transformed Beta variables, ii) an extension of the DM approach to more than 2 variables and, iii) the potential for bias in means or variances of samples produced by the DM approach. These aims are addressed in each of the following sections.

### 3.1. Efficient implementation

The method and template provided by Ren et al. requires the means and variances of the logit transformation of the Beta distributions used to characterise uncertainty of each input variable [3]. These were estimated using 5,000 samples from each of the Beta random variables that were subsequently logit transformed. This approach could be slow and cumbersome, particularly for health economic models built using spreadsheet software (i.e., Microsoft Excel). There does exist an analytic solution which allows for a simple and compact implementation of the DM approach. The analytic solutions for the mean and variance of a logit transformed Beta distribution are given by Eq. 1 and Eq. 2.

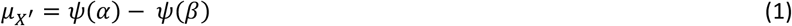

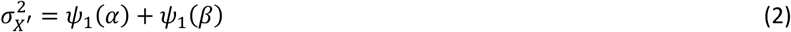

The two parameters of the Beta distribution are *α* and *β, ψ* is the digamma function, *ψ*_1_ is the trigamma function, *X*^′^is the logit transformed Beta random variable and *μ*_*X*_′ and 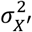are its mean and variance, respectively. The derivations of Eq. 1 and Eq.2 are provided in the Appendix.

For software without built-in functions to approximate the digamma and trigamma functions (e.g., Microsoft Excel), approximations are needed. The expression for the mean in Eq. 1 can be estimated using the asymptotic expansion of the digamma function [5]. The trigamma function is the derivative of the digamma function, an approximation for the variance in Eq. 2 can, therefore, be obtained using the derivative of the asymptotic expansion of the digamma function. The approximations to Eq. 1 and Eq. 2, using the first three terms of each digamma or trigamma approximation, are given in Eq. 3 and Eq. 4.

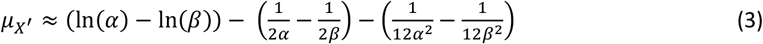

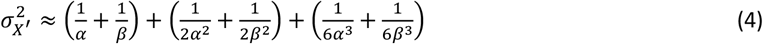

A template in Microsoft Excel that shows how this could be implemented is provided in the Appendix. This template applies the DM approach to a set of five input variables but could be extended to any number of variables (see Section 3.2).

### 3.2. Extension to more than two inputs

When there is a set of more than two variables that are to be sampled with an order constraint, the DM approach requires being able to iterate through the variables, increasing the variance (of the logit transformed beta distributions, 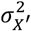) in each step. This involves sampling the variable with the minimum 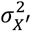and iterating through subsequent variables that are greater than (or less than) this, adding (or subtracting) the sampled differences in each step and thus increasing variance. As an example, consider a set of four variables, *X*_1_,*…,X*_4_, for which 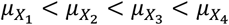 with variances of the corresponding logit transformed beta distributions ordered according to 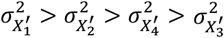. The DM approach could then proceed according to 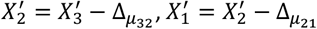and 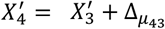

In order to demonstrate the application of the DM approach in more than two variables, health state utilities from two published studies have been examined and are summarised in Table 1. The first are taken from a cost-effectiveness analysis of anti-VEGF agents for the treatment of age-related macular degeneration (AMD) [6]. In this study, health states were defined in terms of visual acuity ranges, and it is assumed that these are ordered such that worse visual acuity must be associated with lower quality of life. The second are taken from an economic evaluation of direct oral anticoagulants in atrial fibrillation [7]. For the purpose of this study, it has again been assumed that these utilities are constrained by the observed order in their point estimates.

**Table 1.** Means and standard errors for two sets of health state utilities, estimated variance of associated logit transformed Beta distributions and errors observed in 10,000 samples using the DM approach SE: standard error; LB: Logit transform of beta distribution with given mean and variance; DM: difference method; VA: visual acuity ^*^The DM error vs target is obtained as the % difference between the simulated results using the DM approach and the target input value, relative to the target ^#^’% X_n_ > X_n+1_’ refers to the proportion of simulated results in which the simulated values using the DM approach exceed the upper bound of the next largest input in the set

| ID | Health state description | Utility | SE | Var(LB) | DM error vs target<sup>1*</sup> | | % $X_n > X_{n+1}$ <sup>#</sup> |
| --- | --- | --- | --- | --- | --- | --- | --- |
|  |  |  |  |  | Mean | Variance |  |
| Case study 1: Age-related macular degeneration |  |  |  |  |  |  |  |
| X<sub>1</sub> | VA: <20/200 | 0.59 | 0.027 | 0.595 | 0.0% | 0.5% | 0% |
| X<sub>2</sub> | VA: 20/50-20/100 | 0.71 | 0.029 | 1.013 | -0.5% | -2.1% | 0% |
| X<sub>3</sub> | VA: 20/30-20/40 | 0.80 | 0.024 | 1.625 | -1.1% | 3.3% | 0% |
| X<sub>4</sub> | VA: 20/20-20/25 | 0.84 | 0.027 | 4.044 | -1.4% | -14.7% | - |
| Case study 2: Atrial fibrillation |  |  |  |  |  |  |  |
| X<sub>1</sub> | Severe stroke sequelae | 0.44 | 0.090 | 0.143 | -0.2% | -2.8% | 0% |
| X<sub>2</sub> | Heart failure | 0.66 | 0.100 | 0.220 | -0.2% | -0.6% | 20% |
| X<sub>3</sub> | Moderate stroke sequelae | 0.75 | 0.040 | 0.047 | -0.1% | 24.8% | 0% |
| X<sub>4</sub> | Atrial fibrillation | 0.78 | 0.005 | 0.001 | 0.0% | 1.3% | - |

The estimated variances of the associated logit beta distributions are given for the two case studies in Table 1. For the first case study in AMD, these variances increase in line with the means of the corresponding utilities. The DM approach can, therefore, iterate through the variables in their natural order. If the variances of the associated logit transformed beta distributions do not monotonically increase in each step from the starting variable at the minimum, then the DM approach will not succeed. This was observed in the second case study, where the minimum variance (of the logit transformed beta distribution) was for *X*_4_ and this variance increased with decreasing utility only until *X*_*2*_. In this situation, two starting values could be used for *X*_*2*_ and *X*_3_, but the order constraint won’t be imposed across the full set – as shown in the final column of Table 1.

Further work to obtain a solution to this issue was considered beyond the scope of the current study. However, it was observed that the variance of a logit transformed Beta random variable as a function of the mean of the Beta random variable is U-shaped. This suggests that the situation observed in the second case study might be unusual, and that a more common situation would be case study 1 or the hypothetical example described at the beginning of this section.

### 3.3. Bias of the difference method approach

One criteria for a successful ordered sampling procedure, as described in Ren et al., is that the summary statistics of the sample should closely match those of the given input variables of interest [3]. When applying the DM approach to the two case studies presented in Table 1, there was evidence of possible bias in the variances of the samples observed in both cases. In the first, the variance of *X*_4_ differed by approximately +15%, and in the second, the variance of *X*_3_ differed by approximately -25%.

To obtain insights into the potential for biased summary statistics when applying the DM approach, a simulation exercise has been performed. This analysis was restricted to two theoretical health state utility values, which were assumed to be bounded by 0 and 1. For a fixed combination of variances, the DM approach was applied over a grid of means for the target variables, obtained by varying each from 0.01 to 0.99 in steps of 0.01. This grid thus consisted of 9,801 unique combinations of input variable means. At each point on the grid, the DM approach was used to produce 100,000 samples for each variable, from which the means and variances were compared to the target values. This was performed for 9 variance combinations that consisted of all possible pairs of 0.002, 0.01 and 0.05.

The resulting errors in the sample means, in terms of a % deviation from the target, are presented in Figure 1 and focus on variable 1 only. This shows that it is possible for the DM approach to produce a sample with a large bias in the sample mean, but only under specific conditions. When a relatively high variance and low mean (variable 1) is combined with a relatively low variance (variable 2), then it is possible to obtain a sample with a mean that is biased downwards. For example, if 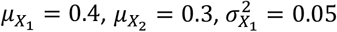and 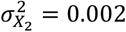, the error in the simulated sample mean for *X*_1_ was -11.9%.

**Fig. 1.**
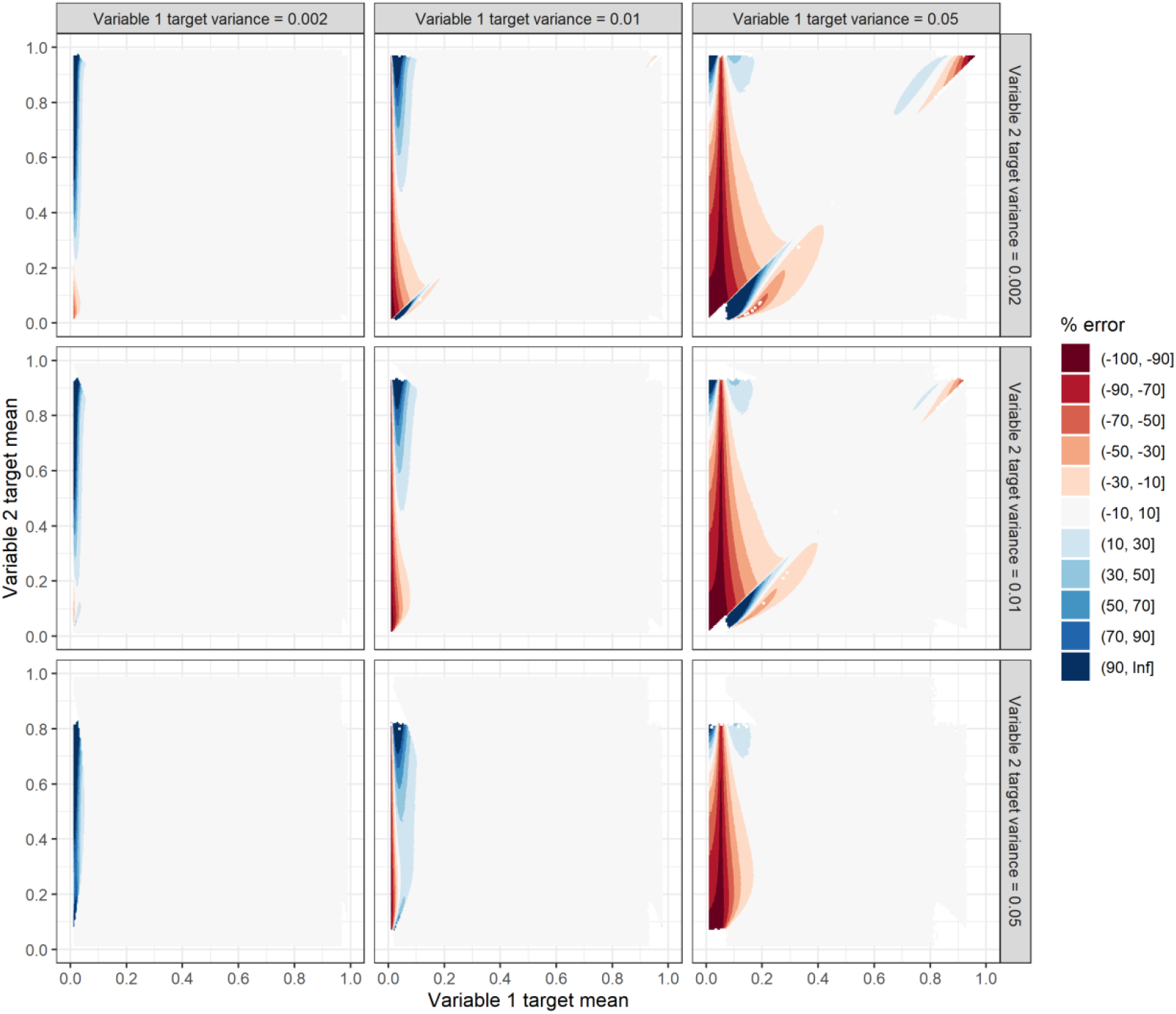
Errors in the sample means obtained via the difference method approach for two variables for varied mean and variance inputs

The resulting errors in the sample variances, in terms of a % deviation compared to the target, are presented in Figure 2, and focus again only on variable 1. This shows that there is a much greater risk that the DM approach will produce a sample with biased variances. It remains the case that this emerges under conditions in which variable 1 has a relatively high variance and this is then constrained by a relatively low variance on variable 2 and/or the lower or upper bound. For example, if 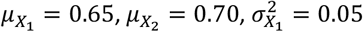and 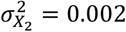, the error in the sample mean for *X*_1_ was -95.4%.

**Fig. 2.**
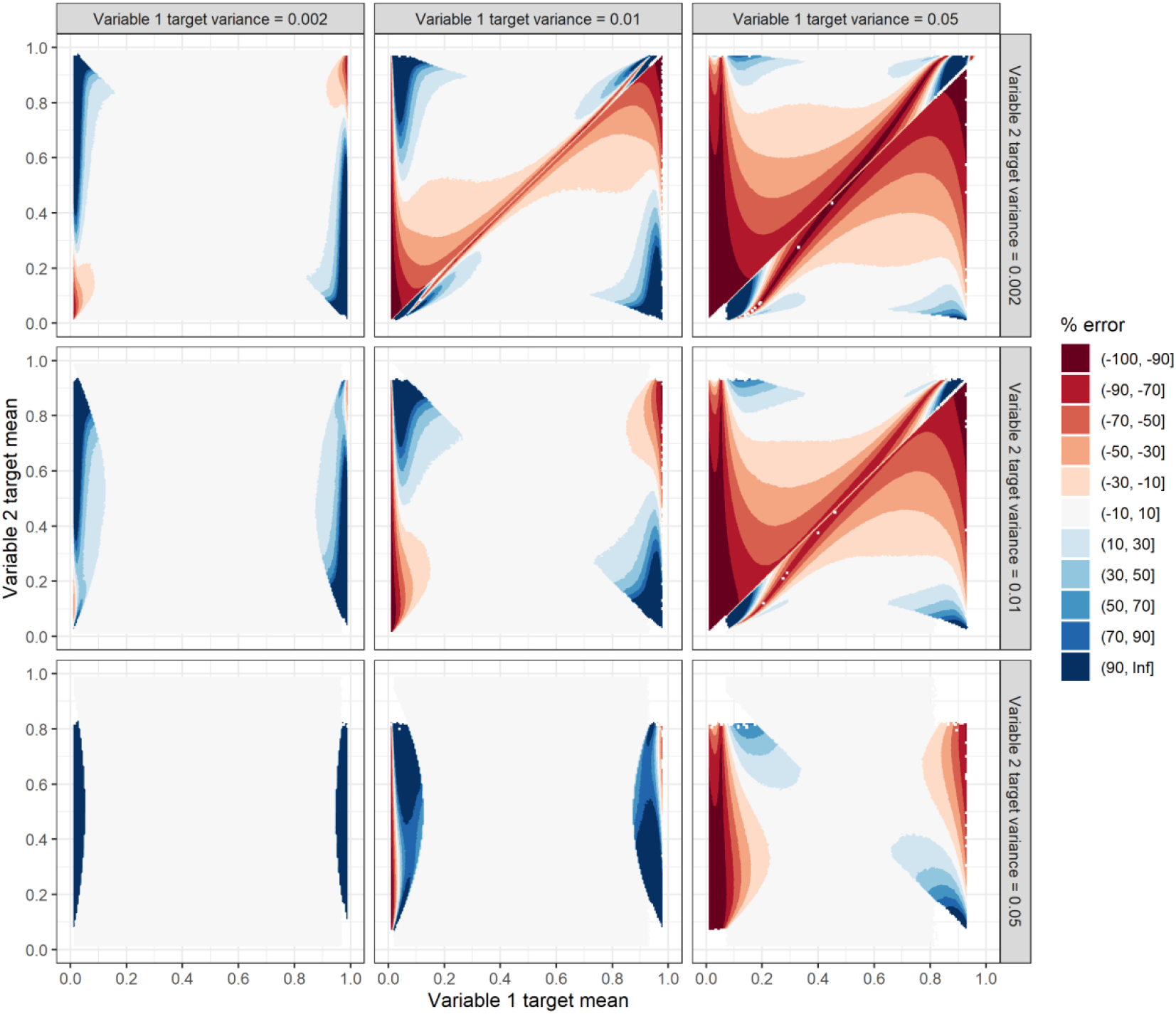
Errors in the sample variances obtained via the difference method approach for two variables for varied mean and variance inputs

## 4. Discussion

This study has developed a simplified implementation of the DM approach [3], that is intended to reduce the technical and computational barriers to its application. It has shown how the method can be extended to an arbitrary number of input variables. However, it has also revealed significant limitations associated with the DM approach. These include situations in which it cannot be fully implemented for a set of more than two variables, and in which it can produce a sample with biased means and variances. As far as the author is aware, these limitations of the DM approach have not been previously reported.

There are several studies that have reported use of the DM approach and cited the original study of Ren et al. [8-10]. These did not report the exact number of variables for which ordered sampling was used, nor did they report on any issues encountered in implementing the approach. It is perhaps noteworthy that none of these studies reported using spreadsheets (i.e., Microsoft Excel) for economic model development. Two were discrete-event simulations, built in R [8] and Python [9], while the third was a Markov model with two living states and choice of software was not reported [10].

Despite the limitations of the DM approach that have been demonstrated in this study, it is possible that the approach will often perform as intended for many model inputs encountered in practice.

Therefore, it is recommended that the DM approach be used for ordered sampling in economic models, unless this is considered unfeasible due to any of these limitations. However, it is important that health economic modellers be aware of its limitations in order to quickly decide on the best approach and avoid biased sampling. To facilitate rapid and straightforward deployment of the DM approach where it is appropriate, for models constructed in Microsoft Excel, a workbook template has been developed and is available at github.com/dh-mcmanus/OS-PSA-template.

Important limitations of the current study include, that no searches were performed for alternative approaches that may be available for ordered sampling of a set of bounded variables. An investigation of alternative probability distributions and the possible implications, as suggested by Ren et al. [3], has not been performed here. This study did not attempt to 1) identify solutions in cases where the DM approach fails for more than two input variables or, 2) to describe the mechanisms that lead to biased means and variances. As regards the situation in which the possibility for negative utilities must be acknowledged, while it was not presented here, this could potentially by handled by first rescaling the original means and variances to the 0,1] interval and then reversing this post-sampling with the DM approach.

## 5. Conclusion

The DM approach for sampling ordered variables has become an established option for health economic modellers implementing probabilistic sensitivity analysis. The increasing awareness of the ordered sampling issue and of the DM approach is expected to lead to an increase in its application. A more straightforward and less computationally intensive implementation of the DM will support its use where appropriate. The DM approach has the limitations that it cannot always be applied to >2 input variables and it can produce biased samples. Knowledge of these limitations can assist the health economic modeller in assessing whether the difference method can be used in a specific application.

## Data Availability

All data produced in the present study are available upon reasonable request to the authors

https://github.com/dh-mcmanus/OS-PSA-template

## 8. Appendix

The article states that the analytic expressions for the mean and variance of a logit transformed Beta distribution are given by Eq. 1 and Eq. 2.

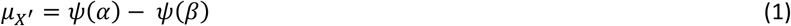

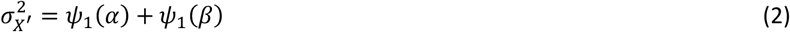

In Eq. 1 and Eq. 2, the two parameters of the Beta distribution are αand β, *ψ*is the digamma function, *ψ*_1_ is the trigamma function, *X*^′^is the logit transformed Beta random variable and *µ*_*X*_^′^and 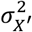are its mean and variance, respectively.

The derivation of Eq. 1 is achieved by writing *E*[*X*^′^]=*E*[*ln*(*X*)−*ln*(1−*X*)]and then evaluating the expected value integrals for a Beta probability density function for each term. For example, for *E*[*ln*(*X*)], the derivation is as follows:

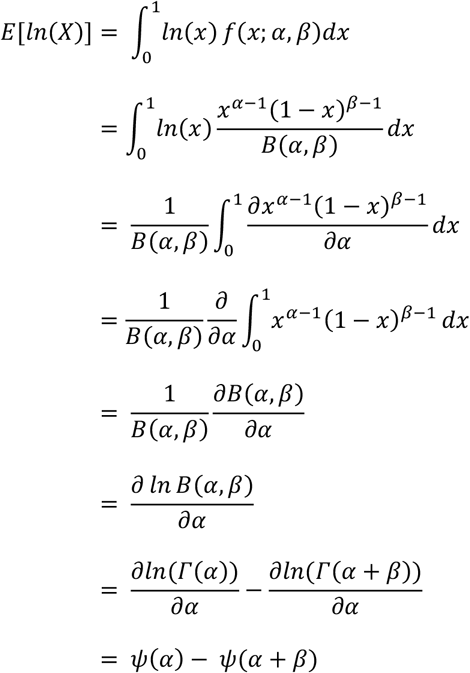

For *E*[*ln*(1−*X*)], substituting *Y*=1−*X*leads, via the same steps, to *ψ*(β)−*ψ*(β+*α*). Therefore, *µ*_*X*_^′^=*E*[*X*^′^]=*ψ*(*α*)−*ψ*(β).

The expression given in Eq. 2 can be obtained by first rewriting the variance of the logit transformed variable as a combination of variances and covariance for *ln*(*X*)and *ln*(1−*X*).

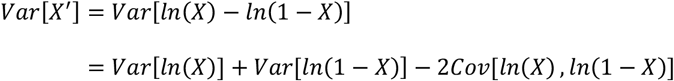

Each of these variance and covariance terms can be obtained using the expressions for the score statistic and Fisher information matrix for a Beta random variable given in Aryal & Nadarajah [11].

The score statistic for the Beta distribution is [11]:

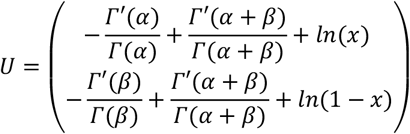

The covariance matrix, or Fisher information matrix, of the score statistic is −*E*[*U*^′^]. The elements of Fisher information matrix are given by [11]:

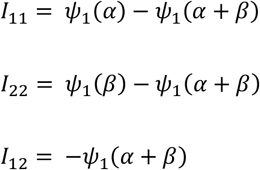

These identities are obtained using the results presented for the translated Beta distribution in Aryal & Nadarajah [11], with *c*=0and *d*=1, and reversing the quotient rule of differentiation.

Since the *Var*[*U*]=−*E*[*U*^′^],it follows that:

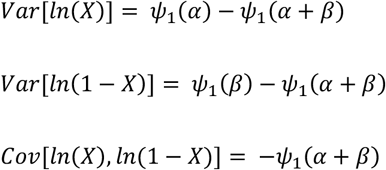

Combining these results with the expression for the variance of *X*^′^given above leads to the result given in Eq. 2.

## Notes

### Competing Interest Statement

The authors have declared no competing interest.

### Funding Statement

This study did not receive any funding

## References

1. Claxton, K., et al., Probabilistic sensitivity analysis for NICE technology assessment: not an optional extra. Health Econ, 2005. 14(4): p. 339–47.

2. National Institute for Health and Care Excellence, NICE health technology evaluations: the manual. 2022.

3. Ren, S., et al., A New Approach for Sampling Ordered Parameters in Probabilistic Sensitivity Analysis. Pharmacoeconomics, 2018. 36(3): p. 341–347.

4. Brazier, J., et al., Identification, Review, and Use of Health State Utilities in Cost-Effectiveness Models: An ISPOR Good Practices for Outcomes Research Task Force Report. Value in Health, 2019. 22(3): p. 267–275.

5. Abramowitz, M. and I.A. Stegun, Handbook of Mathematical Functions with Formulas, Graphs, and Mathematical Tables. 10 ed. 1972: National Bureau of Standards Applied Mathematics Series 55.

6. van Asten, F., et al., The cost-effectiveness of bevacizumab, ranibizumab and aflibercept for the treatment of age-related macular degeneration-A cost-effectiveness analysis from a societal perspective. PLoS One, 2018. 13(5): p. e0197670.

7. Wisloff, T., G. Hagen, and M. Klemp, Economic evaluation of warfarin, dabigatran, rivaroxaban, and apixaban for stroke prevention in atrial fibrillation. Pharmacoeconomics, 2014. 32(6): p. 601–12.

8. Bieri, U., et al., Management of Active Surveillance-Eligible Prostate Cancer during Pretransplantation Workup of Patients with Kidney Failure: A Simulation Study. Clin J Am Soc Nephrol, 2020. 15(6): p. 822–829.

9. Petelin, L., et al., Cost-effectiveness of long-term clinical management of BRCA pathogenic variant carriers. Genet Med, 2020. 22(5): p. 831–839.

10. Thomas, S.A., et al., Behavioural activation therapy for post-stroke depression: the BEADS feasibility RCT. Health Technol Assess, 2019. 23(47): p. 1–176.

11. Aryal, G. and S. Nadarajah, Information matrix for beta distributions. Serdica Mathematical Journal, 2004. 30(4): p. 513–526.

